# HIV incidence among non-migrating persons following a household migration event: a population-based, longitudinal study in Uganda

**DOI:** 10.1101/2023.09.23.23295865

**Authors:** R. Young, J. Ssekasanvu, J. Kagaayi, R. Ssekubugu, G. Kigozi, S.J. Reynolds, M.J. Wawer, B.A.S. Nonyane, Betty Nantume, Thomas C. Quinn, Aaron A.R. Tobian, John Santelli, L.W. Chang, C.E. Kennedy, L. Paina, P.A. Anglewicz, D. Serwadda, F. Nalugoda, M.K. Grabowski

## Abstract

**Background:** The impact of migration on HIV risk among non-migrating household members is poorly understood. We measured HIV incidence among non-migrants living in households with and without migrants in Uganda.

**Methods:** We used four survey rounds of data collected from July 2011–May 2018 from non-migrant participants aged 15–49 years in the Rakai Community Cohort Study, an open, population-based cohort. Non-migrants were individuals with no evidence of migration between surveys or at the prior survey. The primary exposure, household migration, was assessed using census data and defined as ≥1 household member migrating in or out of the house from another community between surveys (∼18 months). Incident HIV cases tested positive following a negative result at the preceding visit. Incidence rate ratios (IRR) with 95% confidence intervals were estimated using Poisson regression with generalized estimating equations and robust standard errors. Analyses were stratified by gender, migration into or out of the household, and the relationship between non-migrants and migrants (i.e., any household migration, spouse, child).

**Findings:** Overall, 11,318 non-migrants (5,674 women) were followed for 37,320 person-years. 28% (6,059/21,370) of non-migrant person-visits had recent migration into or out of the household, and 240 HIV incident cases were identified in non-migrating household members. Overall, non-migrants in migrant households were not at greater risk of acquiring HIV. However, HIV incidence among men was significantly higher when the spouse had recently migrated in (adjIRR:2·12;95%CI:1·05-4·27) or out (adjIRR:4·01;95%CI:2·16-7·44) compared to men with no spousal migration. Women with in- and out-migrant spouses also had higher HIV incidence, but results were not statistically significant.

**Interpretation:** HIV incidence is higher among non-migrating persons with migrant spouses, especially men. Targeted HIV testing and prevention interventions such as pre-exposure prophylaxis could be considered for those with migrant spouses.

**Funding:** National Institutes of Health, US Centers for Disease Control and Prevention

**Research in context:** We searched PubMed for studies focused on HIV acquisition, prevalence or sexual behaviors among non-migrants who lived with migrants in sub-Saharan Africa (SSA) using search terms such as “HIV”, “Emigration and Immigration”, “family”, “spouses”, “household”, “parents”, and “children”. Despite high levels of migration and an established association with HIV risk in SSA, there is limited data on the broader societal impacts of migration on HIV acquisition risk among non-migrant populations directly impacted by it.

There has been only one published study that has previously evaluated impact of migration on HIV incidence among non-migrating persons in sub-Saharan Africa. This study, which exclusively assessed spousal migration, was conducted in Tanzania more than two decades earlier prior to HIV treatment availability and found that non-migrant men with long-term mobile partners were more than four times as likely to acquire HIV compared to men who had partners that were residents. To the best of our knowledge, this is the first study to examine the effect of non-spousal migration, including any household migration and child migration, on HIV incidence among non-migrants

**Added value of this study:** In this study, we used data from the Rakai Community Cohort Study (RCCS), a population-based HIV surveillance cohort to measure the impact of migration on HIV incidence for non-migrant household members. The RCCS captures HIV incident events through regular, repeat HIV testing of participants and migration events through household censuses. Our study adds to the current literature by examining the general effect of migration in the household on HIV incidence in addition to child, and spousal migration. Using data from over 11,000 non-migrant individuals, we found that spousal, but not other types of household migration, substantially increased HIV risk among non-migrants, especially among men. Taken together, our results suggest that spousal migration may be associated with an increased risk of HIV acquisition in the period surrounding and immediately after spousal migration.

**Implications of all the available evidence:** Our findings suggest that spousal migration in or out of the household is associated with greater HIV incidence. Targeted HIV testing and prevention interventions such as pre-exposure prophylaxis could be considered for men with migrant spouses.

## Introduction

Migration is increasingly common in Africa and linked to higher HIV risk.(1,2) Previous studies in sub-Saharan Africa have shown that migrants have a higher risk of HIV acquisition, are more likely to report riskier sexual behaviors, and are less likely to be virally suppressed if living with HIV.(2–4) However, far less is known about how migration impacts HIV risk for non-migrant household members. Here, we theorized that migration could be a broadly disruptive event for the household, changing non-migrants’ relationships, access to resources, and social support, thereby increasing their risk of HIV acquisition.(5) As generalized African HIV epidemics decline,(6,7) identifying population sub-groups at highest HIV risk will be critical for effective targeting of limited HIV prevention resources, such as long-acting injectable HIV pre-exposure prophylaxis.

Migration may impact HIV risk among non-migrating persons through changes to non-migrant residents’ sexual networks and their own individual behaviors. Non-migrants may become more stressed as a result of partner, child, or parent migration. Some individuals may cope with the greater stress, separation of partners, or reduced parental supervision by increasing sexual risk behaviors resulting in higher risk of HIV acquisition.(8–11) Previous studies have shown that non-migrants with migrant spouses are more likely to report riskier sexual behaviors, and acquire HIV or other sexually transmitted infections.(10,12,13) However, most previous studies(14) focus on non-migrant wives whose male spouses migrate for work for long periods of time. Consequently, less is known about how HIV incidence for men is impacted by spousal migration, and if migration impacts non-migrants’ risk of acquiring HIV beyond spousal relationships.

Here, we evaluated whether HIV incidence is higher among non-migrants living in households that migrants enter or leave using data from the Rakai Community Cohort Study, an open population-based HIV surveillance cohort in southcentral Uganda. In addition, we also examined the effect on non-migrants with migrant spouses, and parents with migrant children. Based on existing literature, we hypothesized that non-migrants living in migrant households, especially those with migrant spouses, would be more likely to acquire HIV.

## Methods

### Study overview

We used data collected July 2011-May 2018 from non-migrant participants 15–49 years in the Rakai Community Cohort Study (RCCS), an open, population-based cohort in south-central Uganda. As described elsewhere, the RCCS collects data via a census and survey.(15) The census also records household members’ relationship to head of household and any migration into or out of the household since the last census. The survey collects demographic and self-reported sexual behaviors from residents aged 15-49 years who have been living in the community for at least six months or at least one month with the intention to stay. At time of survey, HIV testing and counselling are also offered with referral to HIV services.

Our analysis included 38 continuously surveyed communities, including 34 inland semi-urban and agrarian with moderate HIV seroprevalence and 4 Lake Victoria fishing communities with extremely high HIV seroprevalence (∼40%) over four survey rounds. The analysis was restricted to HIV seronegative RCCS non-migrant individuals. Non-migrants were those who had reported not moving at two sequential survey visits, allowing for one skipped visit. Individuals who reported moving within communities or returning to communities were classified as mobile and excluded. The RCCS protocol was reviewed and approved by the Uganda National Council for Science and Technology, Uganda Virus Research Institute Research Ethics Committee, Johns Hopkins School of Medicine Institutional Review Board, and Western Institutional Review Board.

### Exposure

The primary exposure was a household migration event, defined as one member moving into (in-migrants) or out of the household (out-migrants) to/from another community between surveys (∼18 months), staying at their destination for at least one month or less than one month with the intention to stay for at least six months. Intention to stay was ascertained by either asking the migrants themselves, the head of household, or a proxy respondent. Migration events were assessed from census and survey data irrespective of age.

Households were initially categorized into three groups based on the direction of migration into the household: (i) no-migration households where no recent migration event was documented; (ii) any household in-migration where member(s) migrated into the household; (iii) any household out-migration where member(s) migrated out. Households with experiencing in- and out-migration in the same visit-interval contributed to both in- and out-analyses.

We further determined the relationship between non-migrants and migrants using their current or previous census household roster. The previous census household roster was used to classify relationships between outmigrants and non-migrant residents. Spousal relationships were defined as long-term consensual unions or marital relationships. We examined relationships where the spouse or child migrated as previous studies suggest that spousal migration may impact sexual behavior and networks for the non-migrant spouse;(10,12,13) and a child migrating may impact the support available for the parent.(16) Four types of relationships between non-migrant and migrants were evaluated: (i) non-migrants with an in-migrating spouse; (ii) non-migrants with an out-migrating spouse; (iii) non-migrant parents with an in-migrating child; and (iv) non-migrant parents with an out-migrating child. Those who experienced both an in and out-migration of their spouse or child in the same visit-interval contributed to both in- and out-analyses.

### Outcome

Our outcome was incident HIV infection, defined as a first positive HIV test preceded by an HIV negative test at the prior visit. Those who missed two consecutive survey rounds were excluded from the analysis. HIV status was determined using a validated testing algorithm which included three-rapid tests and enzyme immunoassays or polymerase chain reaction (PCR) tests to confirm HIV status.(15)

### Statistical methods

We first compared the demographics of men and women included in our analytic sample at baseline to those who were lost to follow-up. We then contrasted the demographics for each visit-interval included in our analysis, comparing any household in-migration to no-migration visit-intervals and any household out-migration to no-migration visit-intervals.

Next, we assessed the association between HIV incidence and migration events occurring during the same visit interval. In these primary analyses, we assumed migration always preceded the HIV incident event. Incidence rate ratios with 95% confidence intervals were estimated using Poisson regressions with generalized estimating equations under an exchangeable correlation structure with robust standard errors.(17) Incident HIV was assumed to occur with equal probability through the visit-interval so, on average, HIV acquisition was assumed to occur at the midpoint of the visit-interval. Regressions were stratified by: (i) gender as biological, social, and structural correlates for HIV seropositivity and viremia differ between men and women;(18,19) (ii) migration into or out of the household; (iii) the relationship between non-migrants and migrants (i.e., any household migration, spouse, child); and, (iv) by community type (inland vs fishing), to account for difference in local HIV epidemic dynamics.(15) To account for differences in demographics across exposed and unexposed groups, we adjusted for likely demographic confounders including five-year age group, education, marital status (for non-spousal regressions), survey round, and residency in an inland or fishing community.

We further assessed how spousal migration and marital status impacted HIV incidence as relationship dissolution is associated with spouses migrating out and relationship formation is associated with spouses in-migrating. We first restricted spousal regressions to those currently married. Then, because marital dissolution is associated with spouses migrating out, we also compared HIV incidence among those who were previously married and had a spouse migrate out compared to those who were previously married but did not have any spousal migration.

Because we were not able to determine if migration preceded or followed acquisition of HIV as both migration and incidence were measured within the same visit-interval, we also evaluated HIV incidence in the visit-interval following spousal migration. For the following visit-interval to be included in this analysis, non-migrants had to remain non-migrants after spousal migration and not experience any in- or out-spousal migration.

Lastly, we evaluated whether sexual behavior changed following in- or out-spousal migration. We compared the sexual behaviors at the start of the visit-interval prior to spousal migration to those at the end of the visit-interval following spousal migration. We classified sexual behaviors into lower and higher risk categories (Supplementary Text S1).

Analyses were conducted using Stata 17 and R version 4.1.1.

## Results

### Overview

Of the 28,085 individuals in the cohort, 20,123 (71.7%) individuals had no recent history of migration observed across the four survey waves (Figure 1) and 13,281 were eligible to be surveyed at more than one study visit. Of those, 1,963 were lost to follow-up and they were more likely to be men, younger, and never married (Supplementary Table S1). In the remaining analytic cohort, 11,318 non-migrant participants (5,674 women) were followed for 37,320 person-years contributing 21,370 visit-intervals. Migration into or out of the household occurred within 28% (6,059/21,370) of visit-intervals. Out of those who contributed three visit-intervals, 52·6% had at least one visit where there was migration into or out of the household (Figure 2**: Longitudinal migration in the household patterns for non-migrant men and women contributing three visit-intervals to the analysis.**Each line represents an individual’s trajectory across the three visit-intervals. HH: household).

**Figure 1:**
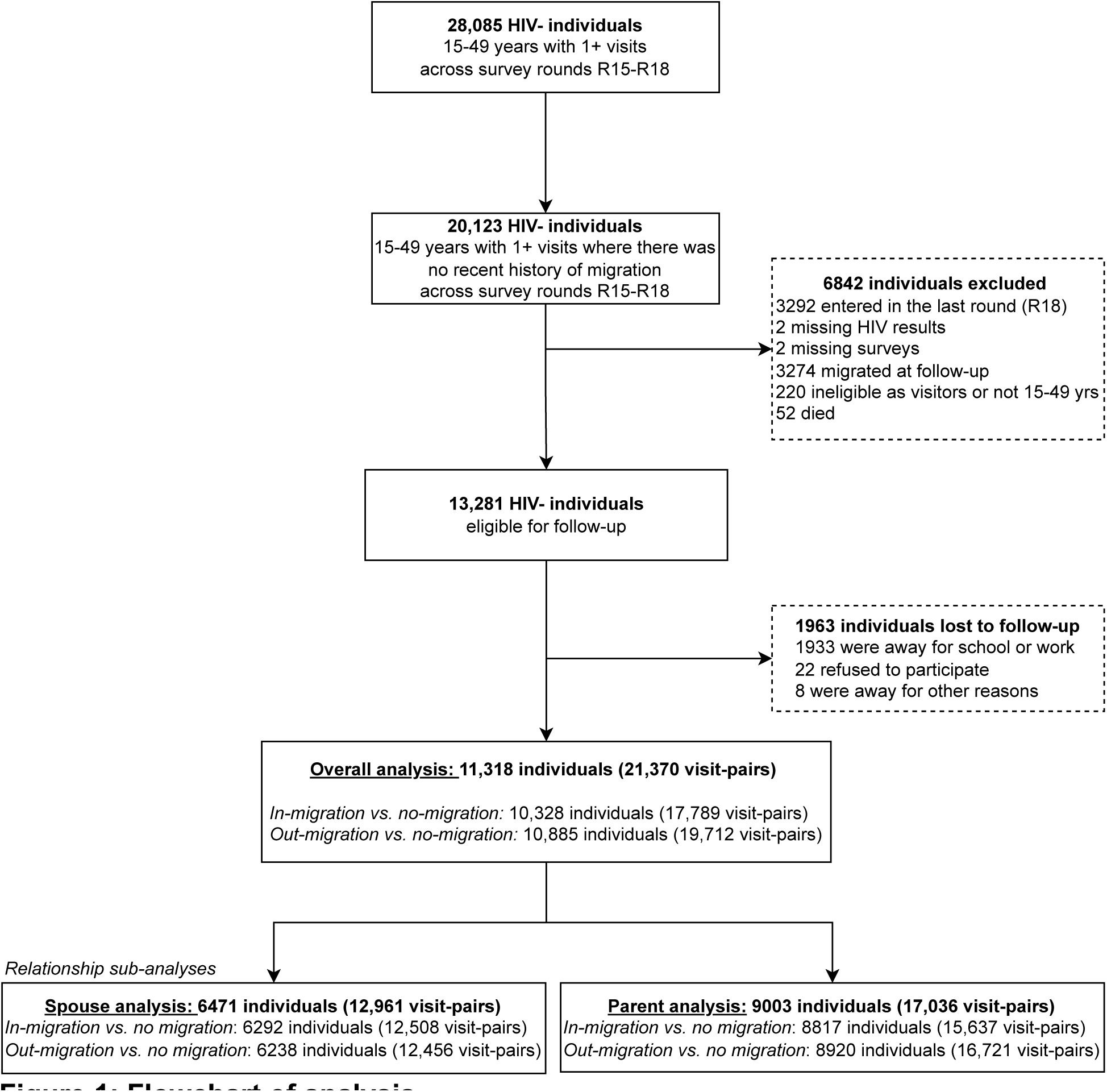
Flowchart of analysis.

**Figure 2:**
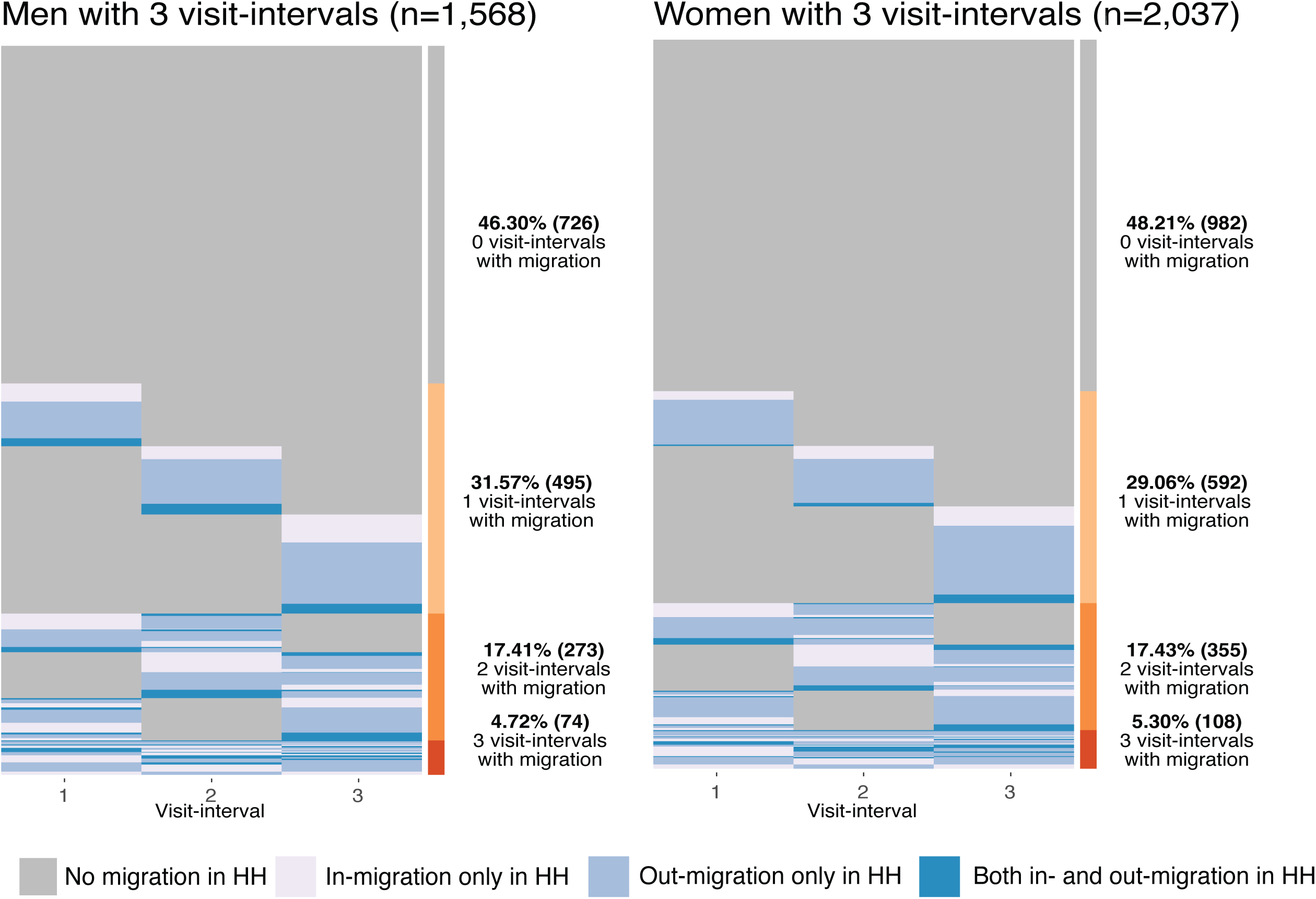
Longitudinal migration in the household patterns for non-migrant men and women contributing three visit-intervals to the analysis. Each line represents an individual’s trajectory across the three visit-intervals. HH: household

At baseline, over half of participants were currently married and most individuals lived in inland communities (Table 1). Visit-intervals from fishing communities were more likely to report migration into the household (Supplementary Table S2). In total 13% (2,820/21,370) of visit-intervals had either a spouse or child migrate. A limited number of visit-intervals (820/21,370; 3·84%) experienced both in- and out-migration in the same period contributing to both in-migration and out-migration households.

**Table 1:** Baseline characteristics of the 11,318 included non-migrant men and women.

|  | **Men** | **Women** |
| --- | --- | --- |
| Individuals | 5644 | 5674 |
| *Education* |  |  |
| None to primary | 3,779 / 5,644 (67%) | 3,445 / 5,674 (61%) |
| Secondary school and beyond | 1,865 / 5,644 (33%) | 2,229 / 5,674 (39%) |
| *Mean age (SD)* | 29 (9) | 30 (9) |
| *Age category* |  |  |
| 15-29 yrs | 2,952 / 5,644 (52%) | 2,809 / 5,674 (50%) |
| 30-39 yrs | 1,678 / 5,644 (30%) | 1,943 / 5,674 (34%) |
| 40-49 yrs | 1,014 / 5,644 (18%) | 922 / 5,674 (16%) |
| *Marital status* |  |  |
| Currently married | 3,030 / 5,644 (54%) | 3,567 / 5,674 (63%) |
| Previously married | 550 / 5,644 (9.7%) | 777 / 5,674 (14%) |
| Never married | 2,064 / 5,644 (37%) | 1,330 / 5,674 (23%) |
| *Community type* |  |  |
| Inland | 4,482 / 5,644 (79%) | 4,894 / 5,674 (86%) |
| Fishing | 1,162 / 5,644 (21%) | 780 / 5,674 (14%) |

Notes: n / N (%); SD standard deviation

### Migration in the household and HIV incidence

There were 240 HIV incident cases identified in non-migrating household members, including 68 (28%) that occurred during the same visit-interval as a household migration event. Overall, there was no association between migration of any household member with risk of HIV acquisition among either men or women (Table 2). Child migration was not associated with increased risk of HIV incidence among parents of either gender.

**Table 2:** Incident rates and ratios for migration in the household.

|  | Exposed |  | Reference<sup>a,b,c</sup> |  | Crude IRR<br>[95% CI] | p | Adj IRR<sup>d</sup><br>[95% CI] | p |
| --- | --- | --- | --- | --- | --- | --- | --- | --- |
|  | Incident<br>cases / pyrs | IR per 100 pyrs<br>[95% CI] | Incident<br>cases / pyrs | IR per 100 pyrs<br>[95% CI] |  |  |  |  |
| Men |  |  |  |  |  |  |  |  |
| Any in-migration<sup>a</sup> | 14/2343 | 0·60<br>[0·35, 1·01] | 77/12655 | 0·61<br>[0·49, 0·76] | 0·98<br>[0·56, 1·74] | 0·95 | 0·85<br>[0·48, 1·5] | 0·58 |
| Any out-migration<sup>a</sup> | 25/3890 | 0·64<br>[0·43, 0·95] | 77/12655 | 0·61<br>[0·49, 0·76] | 1·06<br>[0·67, 1·66] | 0·81 | 1·20<br>[0·77, 1·88] | 0·43 |
| Parent with in-migrating<br>child<sup>b</sup> | 0/181 | .. | 43/11586 | 0·37<br>[0·28, 0·50] | .. | .. | .. | .. |
| Parent with out-migrating<br>child<sup>b</sup> | 2/796 | 0·25<br>[0·06, 1·01] | 43/11586 | 0·37<br>[0·28, 0·50] | 0·68<br>[0·16, 2·80] | 0·59 | 0·79<br>[0·19, 3·26] | 0·74 |
| In-migrating spouse<sup>c</sup> | 13/998 | 1·31<br>[0·76, 2·26] | 34/8534 | 0·40<br>[0·28, 0·56] | **3·27**<br>**[1·73, 6·19]** | **<0·01** | **2·12**<br>**[1·05, 4·27]** | **0·04** |
| Out-migrating spouse<sup>c</sup> | 18/871 | 2·07<br>[1·30, 3·28] | 34/8534 | 0·40<br>[0·28, 0·56] | **5·19**<br>**[2·93, 9·19]** | **<0·01** | **4·01**<br>**[2·16, 7·44]** | **<0·01** |
| Women |  |  |  |  |  |  |  |  |
| Any in-migration<sup>a</sup> | 12/2027 | 0·59<br>[0·34, 1·04] | 95/13895 | 0·68<br>[0·56, 0·84] | 0·87<br>[0·48, 1·58] | 0·64 | 0·88<br>[0·48, 1·62] | 0·67 |
| Any out-migration<sup>a</sup> | 24/3992 | 0·60<br>[0·40, 0·90] | 95/13895 | 0·68<br>[0·56, 0·84] | 0·88<br>[0·56, 1·38] | 0·58 | 0·97<br>[0·61, 1·55] | 0·90 |
| Parent with in-migrating<br>child<sup>b</sup> | 2/405 | 0·49<br>[0·12, 1·98] | 106/15191 | 0·70<br>[0·58, 0·84] | 0·71<br>[0·17, 2·87] | 0·63 | 0·62<br>[0·15, 2·52] | 0·50 |
| Parent with out-migrating<br>child<sup>b</sup> | 6/1754 | 0·34<br>[0·15, 0·76] | 106/15191 | 0·70<br>[0·58, 0·84] | 0·49<br>[0·22, 1·12] | 0·09 | 0·80<br>[0·32, 2·00] | 0·62 |
| In-migrating spouse<sup>c</sup> | 3/228 | 1·32<br>[0·42, 4·10] | 56/12016 | 0·47<br>[0·36, 0·61] | 2·82<br>[0·88, 9·03] | 0·08 | 2·38<br>[0·72, 7·89] | 0·16 |
| Out-migrating spouse<sup>c</sup> | 3/257 | 1·17<br>[0·37, 3·63] | 56/12016 | 0·47<br>[0·36, 0·61] | 2·50<br>[0·78, 8·00] | 0·12 | 2·33<br>[0·72, 7·48] | 0·16 |

**Note:** IR incidence rate; IRR incidence rate ratio; pyrs person-years; CI confidence interval; .. regression did not converge
<sup>a</sup>Compared to no-migration households
<sup>b</sup>Compared to parents with non-migrating children
<sup>c</sup>Compared to spouses with non-migrating spouses
<sup>d</sup>Adjusted for age category, education, study round, fishing or inland community and marital status for non-spouse regressions

However, in analyses of spousal relationships, we observed substantially higher HIV incidence among those with migrating spouses, especially men. Among men who had a spouse who in-migrated or out-migrated into the household, HIV incidence was 1.3 and 2.1 per 100 person-years respectively, compared to 0.4 per 100 person-years among men whose wives had not migrated. Men with an in-migrating spouse were 2·1-fold (95%CI:1·05-4·27) more likely to acquire HIV and men with out-migrating spouses were 4·0-fold more likely (95%CI:2·16-7·44) to acquire HIV compared to men whose spouses did not migrate. Among all non-migrant men, 23% of HIV incident cases (26/111) were detected among men with migrating spouses. HIV incidence was also elevated among women whose spouses migrated into or out of the household compared to women without migrating spouses; however, differences were not statistically significant. To assess whether primary results were sensitive to the assumed timing of HIV acquisition, we ran our primary regressions again but assumed that HIV was acquired at the end of a visit-interval, and this did not alter our findings (Supplementary Table S3).

### Impact of migration in inland and fishing communities

Next, we assessed if analyses by type of community had an effect on the relationship between household migration and HIV incidence. Within inland communities only, men with in- and out-migrant spouses were more likely to acquire HIV compared to men without migrant spouses (Supplementary Table S4). For women in fishing communities, those who experienced any in-migration into the household were 2·28-fold more likely to acquire HIV than women in no-migration households in adjusted analyses (95%CI:1·04-5·01, Supplementary Table S5).

### Marital status and HIV incidence

Examination of spousal migration events showed that 80% (1086/1352) were associated with the breakdown of a relationship or start of a new relationship. To assess whether change in marital status was potentially driving the association between spousal migration and HIV incidence, we stratified analyses by the current marital status of the non-migrant (Supplementary Table S6). Among currently married men, HIV incidence was higher for men whose spouse had in-migrated (adjIRR=1·77; 95%CI:0.76-4.01) but not out-migrated compared to currently married men who did not experience a spousal migration. Men who reported being previously married at the time of survey and who reported spouse out-migration during the visit-interval, were three times as likely to acquire HIV than previously married men who did not experience spousal migration after adjusting for demographics (3.20/100 pyrs vs. 1.23/100 pyrs; adjIRR: 2·95; 95%CI:1·44-6·04).

### Effect of spousal migration on HIV incidence in the visit-interval following spousal migration

Next, we evaluated whether spousal migration in the visit-interval preceding HIV incidence was associated with higher risk of HIV acquisition, as the timing of migration and HIV acquisition events occurring during the same visit-interval was unknown, and HIV incidence may cause spousal migration (Figure 3, Supplementary Table S7). For men, the HIV incidence rate in the visit-interval following spousal migration was 1·78 per 100 person-years (95%CI:0·57-5·57). While this was lower than the estimated incident rate when spousal migration and HIV incidence occurred during same visit-interval (2·39 per 100 person-years (95%CI:1·39-4·12)) it was still substantially higher than that among men whose spouses did not migrate (0·41 per 100 person-years (95%CI:0·29-0·58)). We were unable to assess HIV incidence among men whose spouse migrated into the household in the prior visit-interval as there were no subsequent incident cases. For women, HIV incidence rates in the visit-interval following spousal migration into or out of the household were higher than incidence rates where no spousal migration occurred (following spousal in-migration: 2·44/100 pyrs (95%CI:0·60-9·83); out-migration: 3·51/100 pyrs (95%CI:1·12-11·00)).

**Figure 3:**
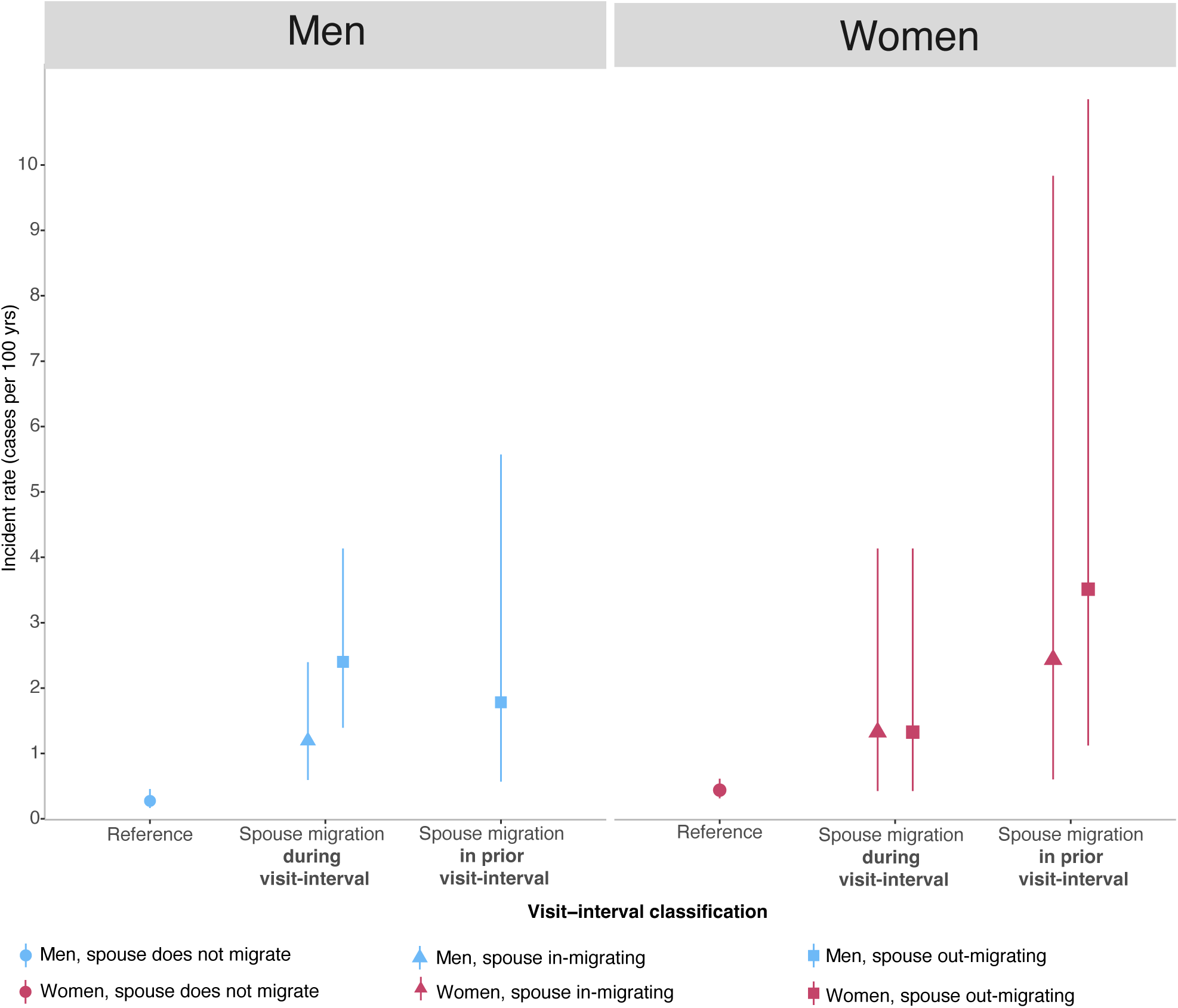
HIV incidence rates and 95% confidence intervals during a spousal migration and following a spousal migration. <u>Note:</u> The incidence rate for men with in-migrating spouses in the prior visit-interval were not able to be estimated as there were zero incident cases. Reference refers to those with spouses that did not migrate.

### Impact of spousal migration on sexual behavior

Lastly, as spousal migration was associated with higher HIV incidence, we compared sexual behaviors prior to spousal migration (t_i-k_) to after spousal migration at the end of the visit-interval (t_i_)(Supplementary Figure S1). Spousal out-migration was associated with an increase in inconsistent condom use with casual partners for 28% of men and 27% of women. For men but not women, spousal migration was also associated with an increased number of sexual partners in the past year (Supplementary Figure S1).

## Discussion

In this population-based study, we found that spousal migration, but not other types of household migration, substantially increased HIV risk among non-migrating household members, especially among men. HIV incidence rates remained elevated among men experiencing spousal migration irrespective of marital status and in the visit-interval following spousal migration, suggesting spousal migration may lead to HIV acquisition. Furthermore, changes in relationship status tended to accompany increases in risk associated with spousal migration. Taken together, our results suggests that spousal migration may be associated with an increased risk of HIV acquisition.

There has been little research on the effect of migration on HIV incidence for non-migrants living with them. To the best of our knowledge, this is the first study to examine the effect of any household migration and child migration on HIV incidence for non-migrants. Only one 2006 study from sub-Saharan Africa examined HIV incidence, using data from couples in Tanzania but this used data from collected between 1994 and 2000 prior to scale-up of HIV treatment and services. Consistent with our findings, they found that non-migrant men with long-term mobile partners were 4·22-fold more likely to acquire HIV compared to men where both partners were residents.(10) Several studies suggest that relationship dissolution, which is associated with spousal migration, is associated with higher rates of incident HIV.(20–22) Studies in the Rakai region also have shown that previously married men and women are more likely to acquire HIV,(20) and that those who are living with HIV are more likely to experience marital dissolution.(23) We also found that that those whose wives had migrated recently were significantly more likely to acquire HIV. This suggests that previously married men are at greatest risk of acquiring HIV in the lead up to and immediately following spousal migration. Women and men whose spouses migrated into the household were also more likely to acquire HIV. This could be due to an increased likelihood for non-migrants, especially men, to partner with migrants with untreated HIV.(24) Our results corroborate with other studies from SSA, demonstrating that changes in sexual behavior following spousal migration are common for men and women with non-migrants reporting riskier sexual behaviors such as reduced condom use and an increased number of sexual partners.(10,11,13)

Our results suggest that non-migrants with migrant spouses may benefit from more focused HIV prevention strategies that mitigate the temporary increase in risk of acquiring HIV during periods surrounding spousal migration. In particular, the use of pre-exposure prophylaxis (PrEP) may benefit non-migrants, especially men, whose spouses have migrated in or out recently. Risk-screening questionnaires are seen as an essential part of making sure that PrEP is able to be accessed by those who need it most.(25) Our findings suggest that such screening tools could include questions on individuals whose spouses have recently migrated. However, further validation and analysis are required to see if screening based on spousal migration is effective.

Furthermore, for non-migrants whose spouses migrate into the household, couple-based HIV interventions could improve HIV outcomes for both the non-migrant and migrant. Previous research has suggested that couple-based interventions, especially couple-based HIV testing and counselling, are more effective than individual-based interventions.(26) Moreover, couple-based HIV testing and counselling should be offered in conjunction with a range of other testing options as couple-based approaches may also have unintended consequences such as increased stigma, couple conflict, or have limited reach.(27) Couple-based testing and counselling can also provide a range of other health benefits that may reduce HIV acquisition risk including increase uptake of testing for other sexually transmitted infections(26,28) and increased condom use,(28) and are designed to improve communication and negotiation between couples.(28)

Our results further suggest that efforts to prevent and treat HIV among migrants are also likely to be beneficial for non-migrants with whom they partner. Migrants are more likely to experience disruptions to HIV treatment due to the challenges associated with migration including reduced time and money and additional structural barriers that make it difficult to transfer care between facilities.(29,30) Several studies suggest that providing testing and treatment outside of health facilities within communities, for extended hours, or that reduce the frequency of refills for ART by dispensing larger amounts may be especially useful for migrants.(29,30) In SSA, previous studies evaluating interventions designed to improve HIV treatment and prevention among migrants are limited as most previous studies highlight barriers in accessing care.(31) However, the interim qualitative findings from an ongoing community-randomized controlled trial designed to link new in-migrants directly to HIV treatment and prevention through dedicated community health workers(32) suggest that community health workers are key in helping migrants access local HIV prevention and treatment services. Beyond community-based care, couple-based interventions to improve ART adherence may also be helpful. Although there are limited studies in Africa, findings suggest that improving communication between couples can improve adherence to ART medications even if one partner is seronegative.(33) In serodiscordant couples where the migrant is living with HIV, maintaining viral suppression will drastically reduce the risk of their non-migrant HIV negative partner from acquiring HIV.

There were several limitations to this study. First, the relative timing of migration into or out of the household and incident HIV was unclear in primary analyses as both were measured during the same survey. However, in secondary analyses, HIV incidence was still elevated in the visit-interval following spousal migration. Second, sexual behaviors were self-reported and may have been underreported. Third, our analyses only go up to 2018. Since 2018, combination HIV interventions have been further scaled up, and so it is unclear if we would observe similar relationships with more recent data. In addition, we were not able to control for individual wealth, the HIV status of new sexual partners for non-migrants, especially those with in-migrant spouses, and the number of previous marriages, all of which may impact HIV incidence. We excluded other types of migration e.g., circular, smaller-scale mobility and return migration, from our analysis. Finally, our analysis is context-specific and as the types, reasons for and broader context for migration varies by setting, our findings may not be generalizable to other settings.

In conclusion, we find that men and women whose spouses migrate into or out of the household are more likely to acquire HIV than those with non-migrant spouses. Our results suggest that HIV prevention interventions should be directed towards individuals with a spouse who recently moved into or out of the household.

## DECLARATIONS

### Contributors

RY and MKG conceived and designed the study. RY, MKG, JSs, JK, RS, GK, SJR, MJW, BAS, BN, TCQ, AART, JSa, LWC, CEK, LP, PAA, DS, and FN contributed to the acquisition, analysis, and interpretation of the data for the study. RY wrote the initial draft manuscript. RY, JS, JK, RS, GK, SJR, MJW, BAS, BN, TCQ, AART, JSa, LWC, CEK, LP, PAA, DS, FN, and MKG contributed to the draft of the manuscript, provided substantial inputs, critical comments and suggested additional analyses. RY and MKG finalized the manuscript. All authors read and approved the final manuscript.

### Availability of data

A de-identified version of the data can be provided to interested parties subject to completion of the Rakai Health Sciences Program data request form and signing of a Data Transfer Agreement. Inquiries should be directed to.

## Acknowledgements

We thank the cohort participants and many staff and investigators who made this study possible. This work was presented as an e-poster at AIDS 2022 and part of RY’s doctoral dissertation. This work was supported by grants from the National Institute of Allergy and Infectious Diseases (grant numbers R01AI114438, R01AI110324, R01AI123002, R01AI128779, R01AI143333, R01AI155080, R01AI087409, U01AI075115, U01AI100031, K25AI114461, K01AI125086), the National Institute of Mental Health (grant number R01MH107275, R01MH105313, R01MH099733, F31MH095649), the Eunice Kennedy Shriver National Institute of Child Health and Human Development (grant number R01HD091003, R01HD050180, R01HD070769), the National Institute on Alcohol Abuse and Alcoholism (K01AA024068),the Division of Intramural Research of the National Institute for Allergy and Infectious Diseases, the Bill and Melinda Gates Foundation (grant number OPP1175094), and the President’s Emergency Plan for AIDS Relief through the Centers for Disease Control and Prevention (grant number NU2GGH000817). RY was supported by the Eunice Kennedy Shriver National Institute of Child Health and Human Development of the National Institutes of Health (grant number 1F31HD102287). We thank the personnel at the Office of Cyberinfrastructure and Computational Biology at the National Institute of Allergy and Infectious Diseases for data management support. The content is solely the responsibility of the authors and does not necessarily represent the official views of the National Institutes of Health.

