## Supplementary Material for "HIV incidence among non-migrating persons following a household migration event: a population-based, longitudinal study in Uganda"

#### Contents


| Supplementary Text | |
|---|---|
| Text S1. Methods to assess changes in sexual behavior following spousal migration | 2 |

| Supplementary Tables | |
|---|---|
| Table S1. Characteristics of those lost to follow-up compared to characteristics of those included at baseline | 3 |
| Table S2. Demographic characteristics of 21,370 non-migrant visit-intervals by household migration: into, out of or no-migration. | 4 |
| Table S3. HIV incidence rate ratios with 95% confidence intervals for household migration and spousal migration assuming HIV is acquired at the end of the visit-interval. | 5 |
| Table S4. HIV incidence for household migration stratified by fishing and inland communities for men | 6 |
| Table S5. HIV incidence for household migration stratified by fishing and inland communities for women | 7 |
| Table S6. HIV incidence and spousal migration by marital status and direction of spousal migration. | 8 |
| Table S7. HIV incidence rates with 95% confidence intervals where spousal migration occurs during the visit-interval, and in the visit-interval following spousal migration | 9 |

| Supplementary Figures | |
|---|---|
| Figure S1. Change (n(%)) in sexual behavior following spousal migration among non-migrant spouses. | 10 |

**Supplementary Text S1. Methods to assess changes in sexual behavior following spousal migration.**

We also evaluated whether sexual behavior changed following in- or out-spousal migration. We compared the sexual behaviors at the start of the visit-interval ($t_{i-k}$) prior to spousal migration to those at the end of the visit-interval ($t_i$) following spousal migration. We restricted our analysis to non-migrant visit-intervals who either had a spouse migrating into or out of the household during that visit-interval and no spousal migration in the prior visit-interval. We excluded those who experienced both in and out-spousal migration in the same visit-interval. Sexual behavior variables were categorized into lower and higher risk categories: condom use (lower risk: consistent use with casual partners or stable partners only; higher risk: inconsistent condom use with casual partners); number of sexual partners in the past year (lower risk: 0-1; higher risk: 2 or more); genital ulcers (lower risk: No; higher risk: Yes). Change in sexual behaviors from the start ($t_{i-k}$) to the end of a visit-interval ($t_i$) were classified as: (i) *increased* defined as move from a lower to higher risk category; (ii) *no change* where the behavior was the same throughout; and (iii) *decreased* defined as moving from a higher to lower risk category.

**Supplementary Table S1. Characteristics of those lost to follow-up compared to characteristics of those included at baseline**

| | Included | Lost-to-follow-up | p-value[a] |
|---|---|---|---|
| **Men** | | | |
| Individuals | 5644 | 1085 | |
| *Education* | | | <0·001 |
| None to primary | 3,779 / 5,644 (67%) | 596 / 1,084 (55%) | |
| Secondary school and beyond | 1,865 / 5,644 (33%) | 488 / 1,084 (45%) | |
| Unknown | 0 | 1 | |
| *Mean age(SD)* | 29 (9) | 25 (9) | <0·001 |
| *Age category* | | | <0·001 |
| 15-29 yrs | 2,952 / 5,644 (52%) | 748 / 1,085 (69%) | |
| 30-39 yrs | 1,678 / 5,644 (30%) | 216 / 1,085 (20%) | |
| 40-49 yrs | 1,014 / 5,644 (18%) | 121 / 1,085 (11%) | |
| *Marital status* | | | <0·001 |
| Currently married | 3,030 / 5,644 (54%) | 426 / 1,085 (39%) | |
| Previously married | 550 / 5,644 (9·7%) | 56 / 1,085 (5·2%) | |
| Never married | 2,064 / 5,644 (37%) | 603 / 1,085 (56%) | |
| Unknown | | | |
| *Community type* | | | 0·2 |
| Inland | 4,482 / 5,644 (79%) | 881 / 1,085 (81%) | |
| Fishing | 1,162 / 5,644 (21%) | 204 / 1,085 (19%) | |
| **Women** | | | |
| Individuals | 5674 | 878 | |
| *Education* | | | <0·001 |
| None to primary | 3,445 / 5,674 (61%) | 362 / 878 (41%) | |
| Secondary school and beyond | 2,229 / 5,674 (39%) | 516 / 878 (59%) | |
| Unknown | 0 | 0 | |
| *Mean age (SD)* | 30 (9) | 23 (9) | <0·001 |
| *Age category* | | | <0·001 |
| 15-29 yrs | 2,809 / 5,674 (50%) | 699 / 878 (80%) | |
| 30-39 yrs | 1,943 / 5,674 (34%) | 122 / 878 (14%) | |
| 40-49 yrs | 922 / 5,674 (16%) | 57 / 878 (6·5%) | |
| *Marital status* | | | <0·001 |
| Currently married | 3,567 / 5,674 (63%) | 265 / 877 (30%) | |
| Previously married | 777 / 5,674 (14%) | 60 / 877 (6·8%) | |
| Never married | 1,330 / 5,674 (23%) | 552 / 877 (63%) | |
| Unknown | 0 | 1 | |
| *Community type* | | | 0·9 |
| Inland | 4,894 / 5,674 (86%) | 759 / 878 (86%) | |
| Fishing | 780 / 5,674 (14%) | 119 / 878 (14%) | |

Notes: n / N (%); SD standard deviation; [a] Pearson's Chi-squared test; Wilcoxon rank sum test

**Supplementary Table S2. Demographic characteristics of 21,370 non-migrant visit-intervals by household migration: in, out of or no-migration.**

| Characteristic | No-migration | In-migration | p-value[1a] | Out-migration | p-value[1b] |
|---|---|---|---|---|---|
| Men | | | | | |
| Visit-intervals | 7,262 | 1,339 | | 2,171 | |
| Person-years | 12,655 | 2,343 | | 3,890 | |
| Survey round[c] | | | 0·8 | | 0·005 |
| R016 (2013-2015) | 2,224 (31%) | 418 (31%) | | 602 (28%) | |
| R017 (2015-2016) | 2,497 (34%) | 449 (34%) | | 733 (34%) | |
| R018 (2016-2018) | 2,541 (35%) | 472 (35%) | | 836 (39%) | |
| Education | | | 0·2 | | 0·004 |
| None to primary | 5,027 (69%) | 905 (68%) | | 1,432 (66%) | |
| Senior school or beyond | 2,235 (31%) | 434 (32%) | | 739 (34%) | |
| Mean age (SD) | 32 (9) | 31 (9) | <0·001 | 30 (11) | <0·001 |
| Age category | | | <0·001 | | <0·001 |
| 15-29 yrs | 2,886 (40%) | 652 (49%) | | 1,127 (52%) | |
| 30-39 yrs | 2,676 (37%) | 397 (30%) | | 473 (22%) | |
| 40-49 yrs | 1,700 (23%) | 290 (22%) | | 571 (26%) | |
| Marital status | | | <0·001 | | <0·001 |
| Currently married | 4,517 (62%) | 941 (70%) | | 957 (44%) | |
| Previously married | 777 (11%) | 73 (5·5%) | | 308 (14%) | |
| Never married | 1,968 (27%) | 325 (24%) | | 906 (42%) | |
| Community type | | | <0·001 | | <0·001 |
| Inland | 5,716 (79%) | 952 (71%) | | 1,836 (85%) | |
| Fishing | 1,546 (21%) | 387 (29%) | | 335 (15%) | |
| Women | | | | | |
| Visit-intervals | 8,049 | 1,139 | | 2,230 | |
| Person-years | 13,895 | 2,027 | | 3,992 | |
| Survey round[c] | | | 0·4 | | <0·001 |
| R016 (2013-2015) | 2,536 (32%) | 337 (30%) | | 621 (28%) | |
| R017 (2015-2016) | 2,729 (34%) | 397 (35%) | | 742 (33%) | |
| R018 (2016-2018) | 2,784 (35%) | 405 (36%) | | 867 (39%) | |
| Education | | | <0·001 | | 0·2 |
| None to primary | 5,124 (64%) | 605 (53%) | | 1,388 (62%) | |
| Senior school or beyond | 2,925 (36%) | 534 (47%) | | 842 (38%) | |
| Mean age (SD) | 32 (8) | 33 (9) | <0·001 | 34 (10) | <0·001 |
| Age category | | | <0·001 | | <0·001 |
| 15-29 yrs | 3,287 (41%) | 363 (32%) | | 669 (30%) | |
| 30-39 yrs | 3,275 (41%) | 489 (43%) | | 732 (33%) | |
| 40-49 yrs | 1,487 (18%) | 287 (25%) | | 829 (37%) | |
| Marital status | | | <0·001 | | <0·001 |
| Currently married | 5,776 (72%) | 727 (64%) | | 1,251 (56%) | |
| Previously married | 1,045 (13%) | 192 (17%) | | 426 (19%) | |
| Never married | 1,228 (15%) | 220 (19%) | | 553 (25%) | |
| Community type | | | <0·001 | | <0·001 |
| Inland | 6,897 (86%) | 925 (81%) | | 2,061 (92%) | |
| Fishing | 1,152 (14%) | 214 (19%) | | 169 (7·6%) | |

Notes: number of visit-intervals (%); SD standard deviation

[1] Pearson's Chi-squared test; Wilcoxon rank sum test

[a] Compares individuals in in-migration households versus no-migration households

[b] Compares individuals in out-migration households versus no-migration household

[c] Second survey of visit-interval, years data was collected in brackets.

**Supplementary Table S3. HIV incidence rate ratios with 95% confidence intervals for household migration and spousal migrations assuming HIV is acquired at the end of the visit-interval.**

| | Crude IRR [95% CI] | p | Adj IRR[c] [95% CI] | p |
|---|---|---|---|---|
| Men | | | | |
| Any household in-migration[a] | 0·98 [0·56, 1·73] | 0·95 | 0·85 [0·48, 1·50] | 0·58 |
| Any household out-migration[a] | 1·05 [0·67, 1·65] | 0·82 | 1·20 [0·77, 1·86] | 0·43 |
| Spouse with in-migrating spouse[b] | 3·24 [1·72, 6·10] | <0·01 | 2·11 [1·06, 4·20] | 0·03 |
| Spouse with out-migrating spouse[b] | 5·10 [2·90, 8·96] | <0·01 | 3·96 [2·16, 7·28] | <0·01 |
| Women | | | | |
| Any household in-migration[a] | 0·87 [0·48, 1·58] | 0·64 | 0·87 [0·48, 1·57] | 0·64 |
| Any household out-migration[a] | 0·88 [0·56, 1·37] | 0·57 | 0·97 [0·61, 1·53] | 0·89 |
| Spouse with in-migrating spouse[b] | 2·79 [0·88, 8·83] | 0·08 | 2·35 [0·72, 7·65] | 0·16 |
| Spouse with out-migrating spouse[b] | 2·48 [0·78, 7·86] | 0·12 | 2·31 [0·73, 7·34] | 0·15 |

Note: IRR incidence rate ratio; CI confidence interval
[a]Compared to no-migration households
[b]Compared to spouses with non-migrating spouses
[c]Adjusted for age category, education, study round, fishing or inland community and marital status for non-spouse regressions

### Supplementary Table S4. HIV incidence for household migration stratified by fishing and inland communities for men

| | Exposed | | Reference[a,b,c] | | | | | |
|---|---|---|---|---|---|---|---|---|
| | Incident cases / pyrs | IR per 100 pyrs [95%CI] | Incident cases / pyrs | IR per 100 pyrs [95%CI] | Crude IRR [95%CI] | p | Adjusted IRR [95%CI][d] | p |
| Men - Fishing | | | | | | | | |
| Any household in-migration[a] | 8 / 610 | 1·31 [0·65, 2·63] | 39 / 2381 | 1·64 [1·19, 2·25] | 0·8 [0·37, 1·72] | 0·57 | 0·84 [0·39, 1·80] | 0·66 |
| Any household out-migration[a] | 9 / 339 | 2·65 [1·38, 5·12] | 28 / 1705 | 1·65 [1·13, 2·40] | 1·15 [0·57, 2·30] | 0·70 | 1·18 [0·59, 2·35] | 0·64 |
| Parent with in-migrating child[b] | 0 / 62 | ·· ·· | 17 / 1463 | 1·16 [0·72, 1·87] | ·· ·· | ·· ·· | ·· ·· | ·· |
| Parent with out-migrating child[b] | 0 / 42 | ·· ·· | 17 / 1463 | 1·16 [0·72, 1·87] | ·· ·· | ·· ·· | ·· ·· | ·· |
| Spouse with in-migrating spouse[c] | 7 / 300 | 2·34 [1·11, 4·92] | 16 / 1243 | 1·29 [0·79, 2·11] | 1·81 [0·75, 4·41] | 0·19 | 1·6 [0·65, 3·92] | 0·30 |
| Spouse with out-migrating spouse[c] | 7 / 253 | 2·77 [1·32, 5·83] | 16 / 1243 | 1·29 [0·79, 2·11] | 2·15 [0·89, 5·22] | 0·09 | 2·28 [0·91, 5·68] | 0·08 |
| Men - Inland | | | | | | | | |
| Any household in-migration[a] | 6 / 1734 | 0·35 [0·16, 0·77] | 38 / 10275 | 0·37 [0·27, 0·51] | 0·94 [0·4, 2·21] | 0·88 | 0·86 [0·36, 2·02] | 0·73 |
| Any household out-migration[a] | 15 / 3358 | 0·45 [0·27, 0·74] | 38 / 10275 | 0·37 [0·27, 0·51] | 1·21 [0·66, 2·2] | 0·54 | 1·24 [0·68, 2·26] | 0·48 |
| Parent with in-migrating child[b] | 0 / 119 | ·· ·· | 26 / 10123 | 0·26 [0·17, 0·38] | ·· ·· | ·· ·· | ·· ·· | ·· |
| Parent with out-migrating child[b] | 2 / 754 | 0·27 [0·07, 1·06] | 26 / 10123 | 0·26 [0·17, 0·38] | 1·03 [0·24, 4·36] | 0·97 | 0·97 [0·23, 4·05] | 0·97 |
| Spouse with in-migrating spouse[c] | 6 / 698 | 0·88 [0·39, 1·95] | 18 / 7291 | 0·25 [0·16, 0·39] | 3·48 [1·38, 8·76] | 0·01 | 2·99 [1·08, 8·23] | 0·04 |
| Spouse with out-migrating spouse[c] | 11 / 618 | 1·79 [0·99, 3·23] | 18 / 7291 | 0·25 [0·16, 0·39] | 7·21 [3·41, 15·28] | <0·01 | 6·53 [3·09, 13·79] | <0·01 |

Note: IR incidence rate; IRR incidence rate ratio; pyrs person-years; CI confidence interval; ·· regression did not converge

[a]Compared to no-migration households

[b]Compared to parents with non-migrating children

[c]Compared to spouses with non-migrating spouses

[d]Adjusted for age category, education, study round, fishing or inland community and marital status for non-spouse regressions

**Supplementary Table S5. HIV incidence for household migration stratified by fishing and inland communities for women**

| | Exposed | | Reference[a,b,c] | | | | | |
|---|---|---|---|---|---|---|---|---|
| | Incident cases / pyrs | IR per 100 pyrs [95%CI] | Incident cases / pyrs | IR per 100 pyrs [95%CI] | Crude IRR [95%CI] | p | Adjusted IRR [95%CI][d] | p |
| Women - Fishing | | | | | | | | |
| Any household in-migration[a] | 9 / 339 | 2·65 [1·38, 5·12] | 28 / 1705 | 1·65 [1·13, 2·40] | 1·61 [0·76, 3·43] | 0·21 | 2·28 [1·04, 5·01] | 0·04 |
| Any household out-migration[a] | 6 / 268 | 2·26 [1·02, 5·04] | 28 / 1705 | 1·65 [1·13, 2·40] | 1·36 [0·56, 3·30] | 0·49 | 1·42 [0·61, 3·32] | 0·41 |
| Parent with in-migrating child[b] | 2 / 103 | 1·95 [0·48, 7·96] | 34 / 1747 | 1·95 [1·39, 2·74] | 1·00 [0·24, 4·20] | 1·00 | 1·65 [0·37, 7·49] | 0·51 |
| Parent with out-migrating child[b] | 1 / 76 | 1·32 [0·18, 9·61] | 34 / 1747 | 1·95 [1·39, 2·74] | 0·68 [0·09, 4·99] | 0·70 | 1·37 [0·21, 9·00] | 0·74 |
| Spouse with in-migrating spouse[c] | 1 / 66 | ·· ·· | 18 / 1423 | 1·27 [0·80, 2·02] | 1·19 [0·16, 9·04] | 0·86 | 1·45 [0·20, 10·46] | 0·71 |
| Spouse with out-migrating spouse[c] | 1 / 52 | 1·92 [0·26, 14·28] | 18 / 1423 | 1·27 [0·80, 2·02] | 1·51 [0·20, 11·58] | 0·69 | 1·75 [0·23, 13·44] | 0·59 |
| Women - Inland | | | | | | | | |
| Any household in-migration[a] | 3 / 1688 | 0·18 [0·06, 0·55] | 67 / 12190 | 0·55 [0·43, 0·70] | 0·32 [0·10, 1·03] | 0·06 | 0·33 [0·10, 1·08] | 0·07 |
| Any household out-migration[a] | 18 / 3724 | 0·48 [0·30, 0·77] | 67 / 12190 | 0·55 [0·43, 0·70] | 0·88 [0·52, 1·48] | 0·63 | 0·86 [0·50, 1·47] | 0·57 |
| Parent with in-migrating child[b] | 0 / 302 | ·· ·· | 72 / 13444 | 0·54 [0·43, 0·68] | ·· ·· | ·· ·· | ·· ·· | ·· |
| Parent with out-migrating child[b] | 5 / 1678 | 0·30 [0·12, 0·72] | 72 / 13444 | 0·54 [0·43, 0·68] | 0·56 [0·22, 1·38] | 0·21 | 0·69 [0·24, 1·97] | 0·49 |
| Spouse with in-migrating spouse[c] | 2 / 162 | 1·24 [0·31, 4·98] | 38 / 10593 | 0·36 [0·26, 0·49] | 3·44 [0·83, 14·28] | 0·09 | 3·63 [0·86, 15·34] | 0·08 |
| Spouse with out-migrating spouse[c] | 2 / 205 | 0·97 [0·24, 3·92] | 38 / 10593 | 0·36 [0·26, 0·49] | 2·71 [0·65, 11·26] | 0·17 | 2·72 [0·66, 11·20] | 0·17 |

Note: IR incidence rate; IRR incidence rate ratio; pyrs person-years; CI confidence interval; ·· regression did not converge

[a]Compared to no-migration households

[b]Compared to parents with non-migrating children

[c]Compared to spouses with non-migrating spouses

[d]Adjusted for age category, education, study round, fishing or inland community and marital status for non-spouse regressions

**Supplementary Table S6. HIV incidence and spousal migration by marital status and direction of spousal migration.**

| | Exposed | | Reference[a,b] | | | | | |
|---|---|---|---|---|---|---|---|---|
| | Incident cases / pyrs | IR per 100 pyrs | Incident cases / pyrs | IR per 100 pyrs | Crude IRR | p | Adj IRR[c] | p |
| Men | | | | | | | | |
| Spouse with in-migrating spouse only[a] | 8/670 | 1·19 [0·60,2·39] | 34/8534 | 0·40 [0·28,0·56] | 3·00 [1·39,6·48] | 0·01 | 1·75 [0·76,4·00] | 0·19 |
| Spouse with out-migrating spouse only[a] | 13/543 | 2·39 [1·39,4·12] | 34/8534 | 0·40 [0·28,0·56] | 6·01 [3·17,11·38] | <0·001 | 4·53 [2·26,9·08] | <0·01 |
| Spouse with both in- and out-migrating spouse[a] | 5/328 | 1·52 [0·63,3·66] | 34/8534 | 0·40 [0·28,0·56] | 3·82 [1·50,9·77] | 0·01 | 2·93 [1·10,7·82] | 0·03 |
| Spouse with in-migrating spouse, among currently married[a] | 8/668 | 1·20 [0·60,2·4] | 34/8532 | 0·40 [0·28,0·56] | 3·00 [1·39,6·50] | 0·01 | 1·75 [0·76,4·01] | 0·19 |
| Spouse with out-migrating spouse, among currently married[a] | 1/167 | 0·60 [0·08,4·32] | 34/8532 | 0·40 [0·28,0·56] | 1·51 [0·21,11·03] | 0·69 | 0·79 [0·11,5·52] | 0·81 |
| Spouse with out-migrating spouse, among previously married[b] | 12/375 | 3·20 [1·82,5·64] | 19/1544 | 1·23 [0·78,1·93] | 2·61 [1·27,5·37] | 0·01 | 2·95 [1·44,6·04] | <0·01 |
| Women | | | | | | | | |
| Spouse with in-migrating spouse only[a] | 3/197 | 1·52 [0·49,4·75] | 56/12016 | 0·47 [0·36,0·61] | 3·27 [1·02,10·46] | 0·05 | 2·68 [0·81,8·91] | 0·11 |
| Spouse with out-migrating spouse only[a] | 3/226 | 1·33 [0·43,4·13] | 56/12016 | 0·47 [0·36,0·61] | 2·85 [0·89,9·11] | 0·08 | 2·62 [0·82,8·4] | 0·11 |
| Spouse with both in- and out-migrating spouse[a] | 0/31 | ·· | 56/12016 | 0·47 [0·36,0·61] | ·· | ·· | ·· | ·· |
| Spouse with in-migrating spouse, among currently married[a] | 3/195 | 1·54 [0·49,4·79] | 56/12010 | 0·47 [0·36,0·61] | 3·30 [1·03,10·54] | 0·04 | 2·68 [0·81,8·9] | 0·11 |
| Spouse with out-migrating spouse, among currently married[a] | 0/35 | ·· | 56/12010 | 0·47 [0·36,0·61] | ·· | ·· | ·· | ·· |
| Spouse with out-migrating spouse, among previously married[b] | 3/191 | 1·57 [0·50,4·90] | 34/2562 | 1·33 [0·95,1·86] | 1·19 [0·36,3·87] | 0·78 | 0·86 [0·24,3·07] | 0·81 |

Note: IR incidence rate; IRR incidence rate ratio; pyrs person-years; CI confidence interval; ·· regression did not converge

[a]Compared to spouses with non-migrating spouses

[b]Compared to non-migrants who were previously married and who did not have any spousal migration

[c]Adjusted for age category, education, study round, fishing or inland community

**Supplementary Table S7. HIV incidence rates with 95% confidence intervals where spousal migration occurs during the visit-interval, and in the visit-interval following spousal migration**

| | Incident cases / pyrs | Incidence rate per 100 pyrs | 95% CI |
|---|---|---|---|
| Men | | | |
| Spouse with no spousal migration (reference) | 15/5457 | 0·27 | [0·17, 0·46] |
| *Spouse migrates in* | | | |
| Spouse migrates in during the visit-interval | 8/670 | 1·19 | [0·60, 2·40] |
| Interval following in-migration of spouse | 0/253 | · | · |
| *Spouse migrates out* | | | |
| Spouse migrates out during visit-interval | 13/543 | 2·40 | [1·39, 4·13] |
| Interval following out-migration of spouse | 3/168 | 1·78 | [0·57, 5·57] |
| Women | | | |
| Spouse with no spousal migration (reference) | 35/7962 | 0·44 | [0·32, 0·61] |
| *Spouse migrates in* | | | |
| Spouse migrates in during visit-interval | 3/197 | 1·33 | [0·43, 4·13] |
| Interval following in-migration of spouse | 2/82 | 2·44 | [0·60, 9·83] |
| *Spouse migrates out* | | | |
| Spouse migrates out during visit-interval | 3/226 | 1·33 | [0·43, 4·13] |
| Interval following out-migration of spouse | 3/85 | 3·51 | [1·12, 11·00] |

Note: CI Confidence interval; pyrs person-years

### Supplementary Figure S1. Change (n(%)) in sexual behavior following spousal migration among non-migrant spouses.

#### A. Men

##### I. Condom use with casual partners

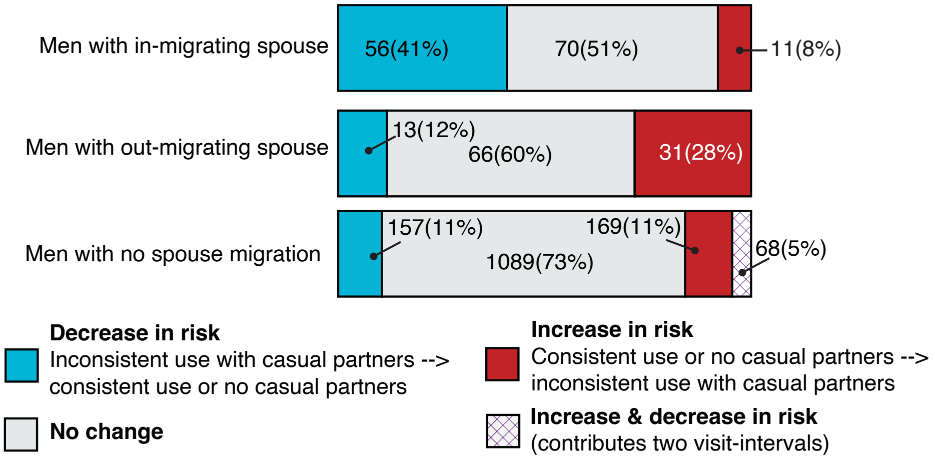


##### II. Number of sexual partners in the past year

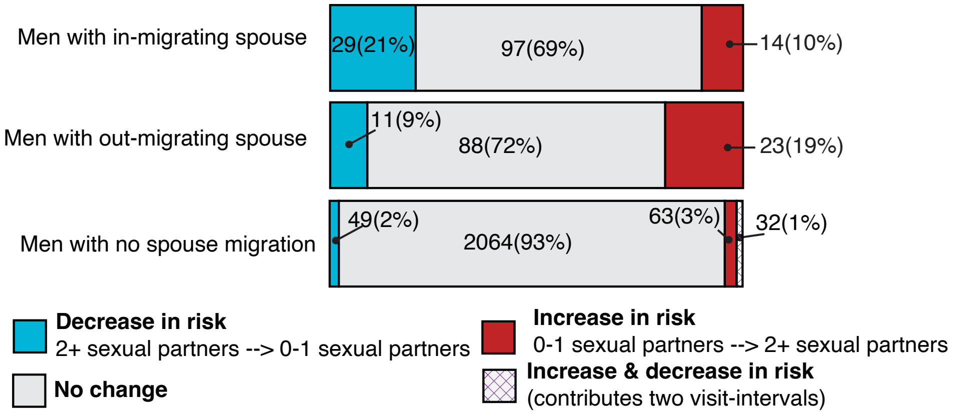


##### III. Genital ulcers

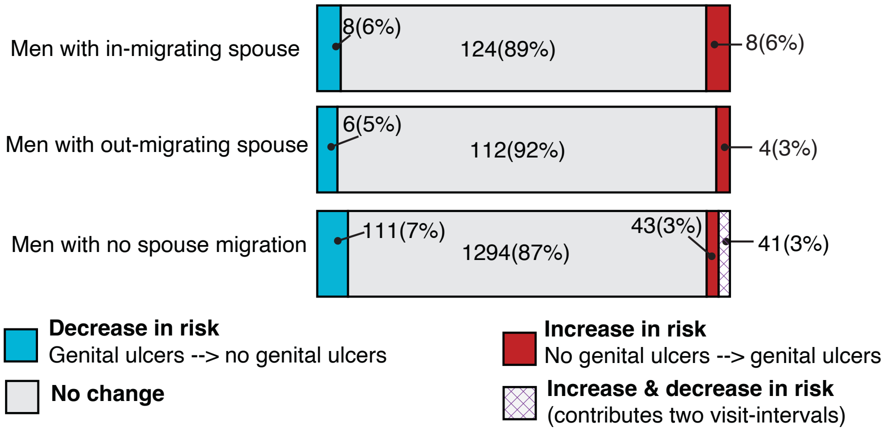

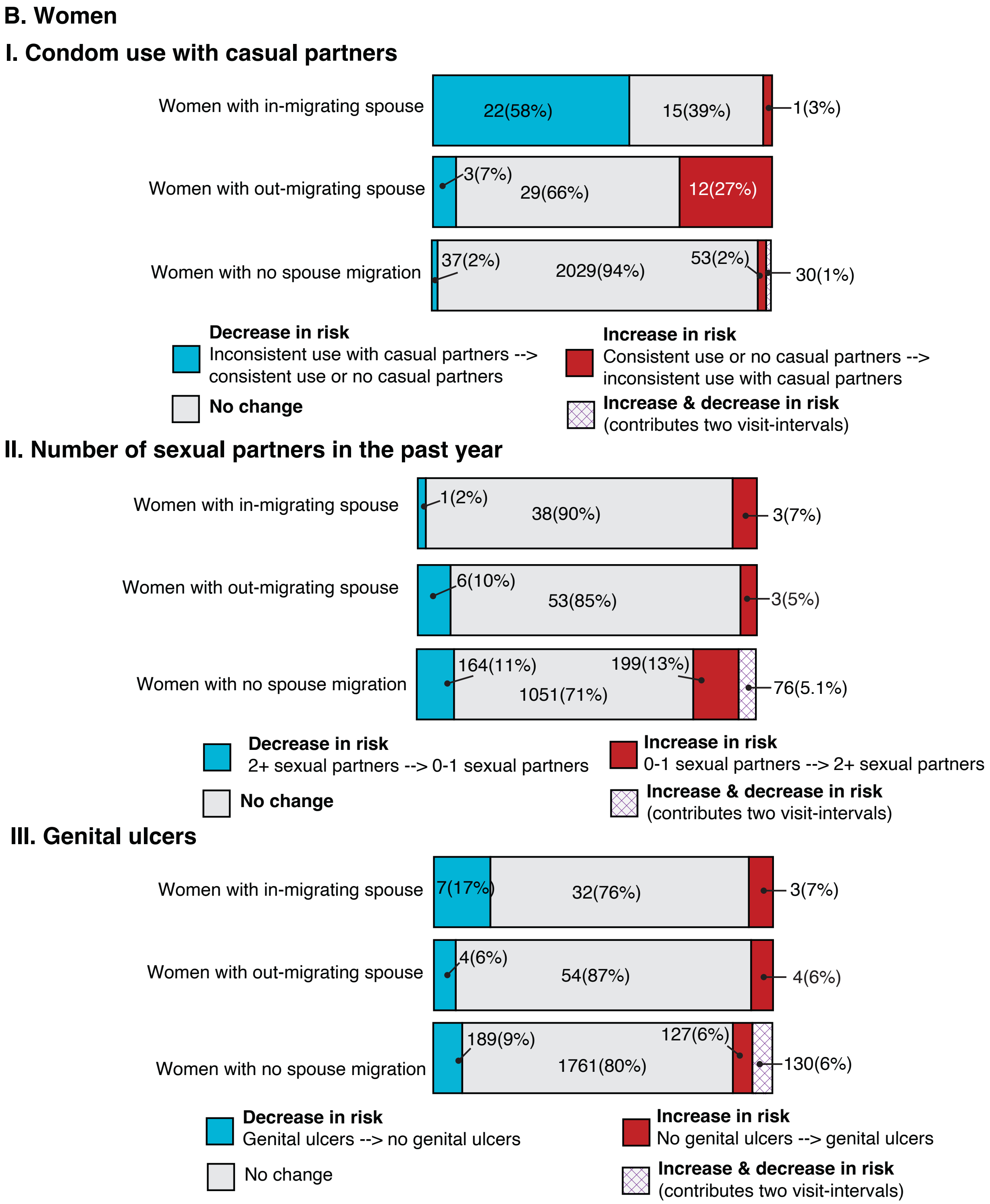
B. Women
I. Condom use with casual partners
Women with in-migrating spouse
22(58%)
15(39%)
1(3%)
Women with out-migrating spouse
3(7%)
29(66%)
12(27%)
Women with no spouse migration
37(2%)
2029(94%)
53(2%)
30(1%)
Decrease in risk
Inconsistent use with casual partners --> consistent use or no casual partners
No change
Increase in risk
Consistent use or no casual partners --> inconsistent use with casual partners
Increase & decrease in risk
(contributes two visit-intervals)
II. Number of sexual partners in the past year
Women with in-migrating spouse
1(2%)
38(90%)
3(7%)
Women with out-migrating spouse
6(10%)
53(85%)
3(5%)
Women with no spouse migration
164(11%)
1051(71%)
199(13%)
76(5.1%)
Decrease in risk
2+ sexual partners --> 0-1 sexual partners
No change
Increase in risk
0-1 sexual partners --> 2+ sexual partners
Increase & decrease in risk
(contributes two visit-intervals)
III. Genital ulcers
Women with in-migrating spouse
7(17%)
32(76%)
3(7%)
Women with out-migrating spouse
4(6%)
54(87%)
4(6%)
Women with no spouse migration
189(9%)
1761(80%)
127(6%)
130(6%)
Decrease in risk
Genital ulcers --> no genital ulcers
No change
Increase in risk
No genital ulcers --> genital ulcers
Increase & decrease in risk
(contributes two visit-intervals)
